# High seroprevalence of SARS-CoV-2 antibodies among people living in precarious situations in Ile de France

**DOI:** 10.1101/2020.10.07.20207795

**Authors:** Thomas Roederer, Bastien Mollo, Charline Vincent, Birgit Nikolay, Augusto Llosa, Robin Nesbitt, Jessica Vanhomwegen, Thierry Rose, François Anna, Corinne Torre, Emilie Fourrey, Erica Simons, Sophie Goyard, Yves Janin, Pierre Charneau, Oxana Vratskikh, Anneliese Coury, Stefan Vanel, Pierre Mendiharat, Klaudia Porten, William Hennequin, Clair Mills, Francisco Luquero

## Abstract

**Background:** A nationwide lockdown was implemented in France on 17 March 2020 to control the COVID-19 pandemic. People living in precarious conditions were relocated by the authorities to emergency shelters, hotels and large venues. Médecins sans Frontières (MSF) then intervened to provide medical care in several of these locations in Paris and in Seine-Saint-Denis, one of its suburbs, between March and June 2020. A seroprevalence survey was conducted to assess the level of exposure to COVID-19 among the population living in the sites. To our knowledge, this is the first assessment of the impact of the pandemic on populations living in insecure conditions in Europe.

**Methods:** We conducted a cross-sectional seroprevalence study in the food distribution sites, emergency shelters and workers residences supported by MSF in Paris and Seine-Saint-Denis, to determine the extent of COVID-19 exposure as determined by SARS-CoV2 antibody seropositivity. The detection of SARS-COV2 antibodies in serum was performed at the Institut Pasteur of Paris using two LuLISA (Luciferase-Linked Immunosorbent Assay) assays and a Pseudo Neutralization Test. A questionnaire covering sociodemographic characteristics, living conditions, adherence to sanitary recommendations and symptom manifestations was also completed. We describe here the seroprevalence site by site and identify the risk factors for seropositivity using a multivariable logistic regression model with site random effects. We also investigated associations between seropositivity and symptoms eventually reported.

**Findings:** Overall, 426/818 individuals tested positive in the 14 sites investigated. Seroprevalence varied significantly with the type of site (chi^2^ p<0.001). It was highest at 88.7% (95%CI 81.8-93.2) among individuals living in workers’ residences, followed by 50.5% (95%CI 46.3-54.7) in emergency shelters and 27.8 % (95%CI 20.8-35.7) among individuals recruited from the food distribution sites. Seroprevalence also varied significantly between sites of the same type. Among other risk factors, the odds for seropositivity were higher among individuals living in crowded sites (medium: adj. OR 2.7, 95%CI 1.5-5.1, p=0.001; high: adj. OR 3.4, 95%CI 1.7-6.9, p<0.001) compared with individuals from low crowding sites and among those who reported transit accommodation in a gymnasium before the lockdown (adj. OR 3.1, 95%CI 1.2-8.1, p=0.023). More than two-thirds of the seropositive individuals (68.3%; 95%CI 64.2-72.2) did not report any symptoms during the recall period.

**Interpretation:** The results demonstrate rather high exposure to SARS-COV-2 with important variations between study sites. Living in crowded conditions was identified as the most important explanatory factor for differences in levels of exposure. This study describes the key factors which determine the risk of exposure and illustrates the importance of identifying populations at high risk of exposure in order to orient and adapt prevention and control strategies to their specific needs.

## Introduction

A novel coronavirus causing a severe respiratory syndrome, severe acute respiratory syndrome coronavirus 2 (SARS-COV-2), emerged at the end of 2019 in Hubei province, China and then spread worldwide (1). Following this, Europe became the major hotspot of the global pandemic (1) and the first confirmed cases of the novel coronavirus disease (COVID-19) were detected in France by 24 January 2020 (2). As a response to an exponential transmission rate, with hospitalizations and deaths doubling every two to three days, a nationwide lockdown was implemented on the 17th of March 2020. Although the lockdown applied to the entire country, important differences were observed between regions in terms of number of confirmed cases reported and deaths. Although the final number of infections is yet to be established in France, a model-based study estimated a nationwide infection rate of 6% in France during the first wave, ranging from 1.5% infected in Nouvelle Aquitaine to 12% in Ile-de-France (IDF) (3). A recent seroprevalence study reported similar estimates: 10% in IDF and 3.1% in Nouvelle Aquitaine.(4)

Heterogeneity in the risk of exposure to COVID-19 likely exists among different subpopulations. Specific subgroups such as health care professionals or nursing home employees and residents have already been identified as groups at higher risk of exposure than the general population (5-8). It is likely that other populations have suffered higher exposure risks to COVID-19 because of their working (9,10) or living conditions (i.e.: shared housing). In France, an estimated 900,000 people lack a permanent housing, with an estimated 250,000 people experiencing recurrent homelessness, including up to 50,000 in Ile-de-France. It has been estimated that at least 3,500 people are homeless on the streets of Paris and close to 7,000 in the Ile-de-France (11,12). People experiencing homelessness or otherwise living in precarious conditions may be particularly vulnerable to exposure to COVID-19. Shared housing, including shelters and encampments, and poor sanitary conditions are factors that can potentially expedite virus transmission. Many of the recommended COVID-19 prevention measures, such as social-distancing and self-isolation if symptomatic, may be challenging or not feasible for a population living under these circumstances. In addition, people experiencing homelessness include older adults who may have underlying medical conditions with higher risks of developing severe COVID-related illness (13,14).

During the nationwide lockdown in France, known as “confinement”, French authorities relocated people experiencing homelessness into emergency shelters, including hotels and large venues, such as gymnasiums. Non-governmental organisations and associations filled several resulting gaps in this relocation, including medical care. Between March and June 2020, the non-profit medical humanitarian organization Médecins sans Frontières (MSF) provided medical care and hygiene promotion to populations living in workers’ residences and emergency shelters, in Ile-de-France (IDF). Mobile clinic activities included clinical management of non-COVID cases, with hospital referrals when needed, COVID screening, and referrals to COVID isolation centres when necessary. The MSF mobile clinics performed some sample collection for PCR-based COVID assays but many individuals with mild symptoms compatible with COVID were never tested and the actual rate of infection among the homeless population in France remains unknown.

Given the variable access to testing, and potentially the substantial proportion of asymptomatic cases who did not seek such tests, the number of confirmed cases of positive PCR assays does not reflect the true impact of the pandemic. Serological-based assays can accurately identify the number of people exposed to the virus (16) but although several serological/seroprevalence studies have been undertaken nation-wide, few have focused on populations experiencing homelessness (3,14,15). Recommendations from the National Health Authority (Haute Autorité de Santé) have underscored the need for more serological surveys in specific populations, including among vulnerable groups, to evaluate the overall exposure to the virus, raise awareness and improve preparation for the rebound in COVID-19 cases (16), currently observed (17). This survey was conducted to assess the level of exposure to COVID-19 among the population living in sites served by MSF in IDF. To our knowledge, this is the first survey to assess the pandemic’s impact on vulnerable populations in Europe.

## Methods

### Survey design and sampling strategy

We conducted a cross-sectional seroprevalence study in sites supported by MSF in Paris and Seine-Saint-Denis, two urban departments of Ile de France region, to determine the extent of COVID-19 infection among the population residing in or frequenting these sites, as determined by SARS-CoV2 antibody seropositivity.

A target sample size of 791 individuals was based on the hypothesis that the seroprevalence of anti-SARS-COV2 antibodies among populations living in insecure conditions served by MSF would be two to three fold higher than the modelled estimate for the general population in IDF of 12% (3).

Recruitment sites were identified among the 39 MSF intervention sites based on feasibility of conducting the survey, i.e. whether the sites were still open at the time of survey, security constraints and the sites managers’ agreement. The number of people per site was calculated in proportion to each of the expected site’s population. Training of surveyors and translators was conducted for two days before the initiation of the study. Individuals were randomly selected for participation at each site, using simple random sampling when a listing of residents existed and systematic random sampling otherwise. To increase participation of those initially selected to participate, sites were visited several times at different times of the day, including on weekends and evenings. In case of refusal or absence, the initially selected person was replaced by the next person in line or another adult sharing the room.

After obtaining written informed consent from the participant, a questionnaire was completed face-to-face by a trained interviewer in the participant’s language (for French, English, Arabic, Farsi, Spanish and Portuguese interviews were conducted in person and other languages using a translator by telephone).

Responses were recorded electronically via a Kobo Collect form on a cell phone or tablet. The data were analysed using Stata V.15 software (StataCorp. 2017. College Station, TX) and R (R 3.6.2).

Blood samples were collected on site from each participant, transported and processed within 24 hours for testing.

### Laboratory Procedures

The serology assays for the detection of SARS-COV2 antibodies were performed using the LuLISA (Luciferase-Linked Immunosorbent Assay) technology designed by Institut Pasteur, Paris (18). The LuLISA uses the nucleoproteins (N) of the SARS-CoV-2 virus, or the Spike (S) protein as target antigens for the detection of specific IgG antibodies in human serum using a variable domain of the IgG single heavy chain from immunized alpaca, specific for human IgG constant domain, expressed as a tandem with a luciferase, NanoKAZ (19). In the presence of luciferase substrate, hikarazine-Q108, the luminescence intensity (relative light unit/s) yielded by luciferase catalytic activity is related to the number of target-bound IgG (19, 20). In addition to LuLISA (N), and LuLISA (S), a Pseudo Neutralization Test (PNT) was performed to confirm the presence of antibodies and to assess their potential to neutralise and protect against the SARS-COV-2 virus.

The LuLISA technique can be used to assess the incidence of all the antibodies involved in a viral infection response (IgA, IgM and, as used here, IgG) and is considered highly sensitive. The incidence of IgG in patients with a COVID-19 positive PCR test sampled 15 days after onset of symptoms is 100% (21). The specificity of the LuLISA varied between 97 and 100% in another recent evaluation (21).

The LuLISA and PNT results are expressed in relative light units per second (RLU/s) because they are bioluminescence-based. Positivity for LULISA assays is defined by a cut-off eliminating at least 98% of pre-pandemic samples (10,291 RLU/s). The pseudo-neutralisation positivity threshold is defined by a cut-off corresponding to 3 times the standard deviation measured with the same technique on pre-pandemic sera (28,532 RLU/s). We considered an individual seropositive if at least one of the 3 tests (LuLISA N, LuLISA S or PNT) gave a positive result.

### Statistical Analysis

We describe the study participants using summary measures and estimated the seroprevalence and 95% confidence intervals (95%CI) by site, type of site, and characteristics of participants using the Clopper–Pearson method. A sensitivity analysis of seroprevalence estimates by type of sites, taking into account assumptions about diagnostic test performances, is presented in the supplementary material.

To investigate seropositivity risk factors, we first performed univariable logistic regression analysis by type of site and for all sites combined. We subsequently constructed a multivariable logistic regression model with random intercepts for specific sites of recruitment to account for clustering of individuals.

In the multivariable model selection, we included variables with the potential to epidemiologically explain the differences in seropositivity incidence. We first grouped variables into four categories: (i) sociodemographic characteristics (sex, age, working before lockdown, language barrier), (ii) frequency of leaving the place of residence during the lockdown (for work, cultural activities, use of public transport, time spent outside), (iii) crowding in the place of residence (number sharing room, number sharing sanitary facilities, number sharing kitchen, and number of contacts inside place of residence per day), and (iv) adhering to hygiene recommendations (hand washing, wearing masks, distancing, cleaning, and following recommendations in general).

For sociodemographic characteristics and adherence to hygiene recommendations, we selected variables from each category that were most strongly associated with seropositivity. To investigate associations of seropositivity with population density in a residence, we created a cumulative crowding indicator, based on the sum of the levels of each of the 4 questions summarizing the crowding information for sharing (i) the bedroom, (ii) the shower, (iii) the kitchen, and (iv) the number of close contacts (>15 minutes per day closer than 1 meter). This made it possible to categorize low, medium, and high levels of crowding (see Supplementary material for more details on how the composite crowding indicator was calculated).

We also constructed a similar score for the frequency of leaving place of residence, combining the answers to the questions ‘Gone out to go to work’, ‘Gone out to go to a distribution/association/church site…’ and ‘Took public transport’ into 3 categories: those who never went out, those who rarely/sometimes went out, and those who went out frequently (every day).

Other variables investigated were: tobacco use, awareness of close contact with a COVID case(s), transit through a gymnasium at the beginning of the confinement (as an indicator for exposure to crowding before moving to a specific site), and the type of site. We then performed backwards variable selection; variables with p<=0.05 were retained in the final model. A sensitivity analysis of the multivariable risk factor analysis is presented in the supplementary material.

### Ethical considerations

This protocol was approved by the MSF ethics review board on 18 June 2020, reference number 2044) and by the Comité de Protection des Personnes (CPP), Ile de France, Paris XI, approved 19 June 2020 (reference number 20050-62628).

## Results

Between 23 June and 2 July 2020, we conducted a cross-sectional seroprevalence study among 829 people living in 14 facilities: 2 food distribution sites, 2 workers’ residences and 10 emergency shelters. Depending on the site, between 1 in 10 and one third of participants randomly selected for participation were replaced with another participant from the same site, due to absence or refusal. After cleaning the data and consolidating the results, 818 people were included in all the sites and received their serological results for these SARS-COV-2 assays.

### Characteristics of study participants

Overall, 79.6% (95%CI 76.7-82.3) of study participants were male with an overall sex ratio of 3.9. There were no females present in the workers’ residences. The mean age of participants was 39 years, with 49.0% (95%CI 45.5-52.5) of the population younger than 35 years; the population recruited from food distribution sites was older on average than the one living in shelters (mean age=31.8 vs 49.0; p=0.001). Comorbidities were reported for 20.8% (95%CI% 18.1-23.7) of participants; the most commonly reported health problems were hypertension (8.3%; 95%CI 6.5-10.4) and diabetes (5.6%; 95%CI 4.1-7.4). More than half of the surveyed population reported never smoking (54.1%; 50.6-57.6).

Participants recruited from food distribution sites resided in different types of housing: 18.2% (95%CI 6.4-41.8) in their own residence, 40.9% (95%CI 21.6-63.5) in shelters (emergency or other) and 36.3% (95%CI 18.2-59.4) in the streets or in a camp.

The majority of participants (90.7%, 95%CI 88.5-92.6) were not born in France, and almost a quarter of all participants (22.7%; 95%CI 19.9-25.8) had been residing in France for less than a year. Two-thirds (68.1%; 95%CI 64.8-71.3) of participants reported having medical coverage (French national social security or French state medical aid); 64.3% (95%CI 60.9-67.6) reported an education level up to middle school. Overall, most participants were not working before the lockdown (77.5 % [74.4-80.3]), with the exception of participants from the workers’ residences, 59.7 % (9%CI 50.5-68.4] of whom reported a professional activity before the lockdown.

Living conditions varied by site type. Workers’ residences were more crowded than the other types, where one third (33.6%, 95%CI 25.3-42.7) of participants reported sharing a room with 2-5 other individuals and 21.3% (95%CI 14.4-29.6) with more than five individuals. Among participants from emergency shelters, 59.3% (95%CI 55.0-63.6) shared a room with one other person but rarely with more than five individuals (4%, 95%CI 2.5-6.1), while nearly half (47.8%, 95%CI 39.2-56.5) of participants recruited from food distribution sites did not share accommodation with anyone.

### Seroprevalence

Overall, 426/818 individuals tested positive by any serological test. Seroprevalence varied significantly by type of site (chi^2^ p<0.001) (Figure 1). It was highest at 88.7% (95%CI 81.8-93.2) among individuals living in the two workers’ residences, followed by 50.5 % (95%CI 46.3-54.7) in the 10 emergency shelters and 27.8 % (95%CI 20.8-35.7) among individuals recruited from the two food distribution sites. Seroprevalence also varied between facilities of the same type: 23-62% in emergency shelters, 18-35% in food distribution sites and 82-94% in workers’ residences.

**Figure 1.**
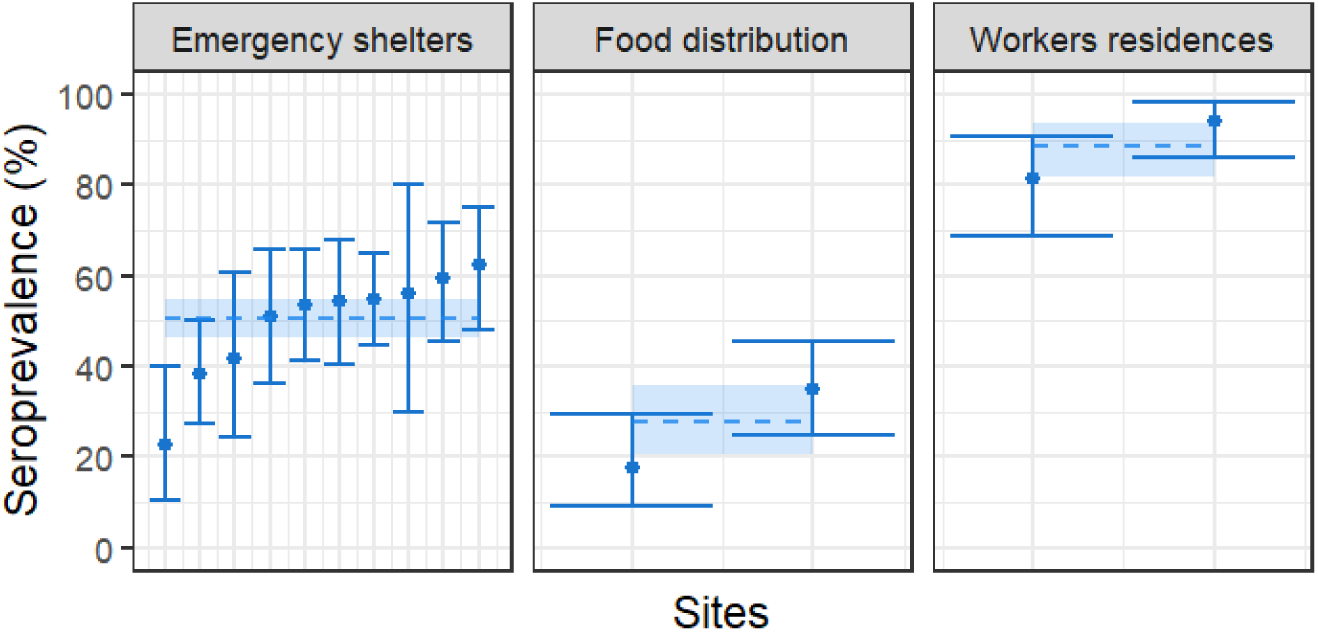
Seroprevalence by type of recruitment site.

The pseudo-neutralization test was positive for 303 out of the 818 individuals. This assay suggests that the seropositive population could therefore be protected against COVID-19 infection, at least at the time of the sample survey (supplementary materials, Table S1).

Correlations and concordance between the three serology techniques were found to be strong (supplementary materials Figure S2-3 and Table S1).

#### Risk factors

In univariable analysis, the most strongly associated risk factors of seropositivity were those linked to crowded living conditions. The odds of seropositivity was 4.3 times (95%CI 2.2-8.4, p<0.001) higher among individuals sharing a room with more than five individuals compared to those not sharing a room; and 3.1 (95%CI 2.0-5.0, p<0.001) times higher among individuals sharing sanitary facilities with more than five individuals compared to those not sharing facilities (Table 1). The odds of seropositivity increased with the level of crowding with an odds ratio (OR) of 3.6 (95%CI 2.0-6.3; p<0.001) for medium and an OR of 6.7 (95%CI 3.6-12.5; p<0.001) for high crowding compared to individuals with a low crowding composite indicator.

**Table 1.**
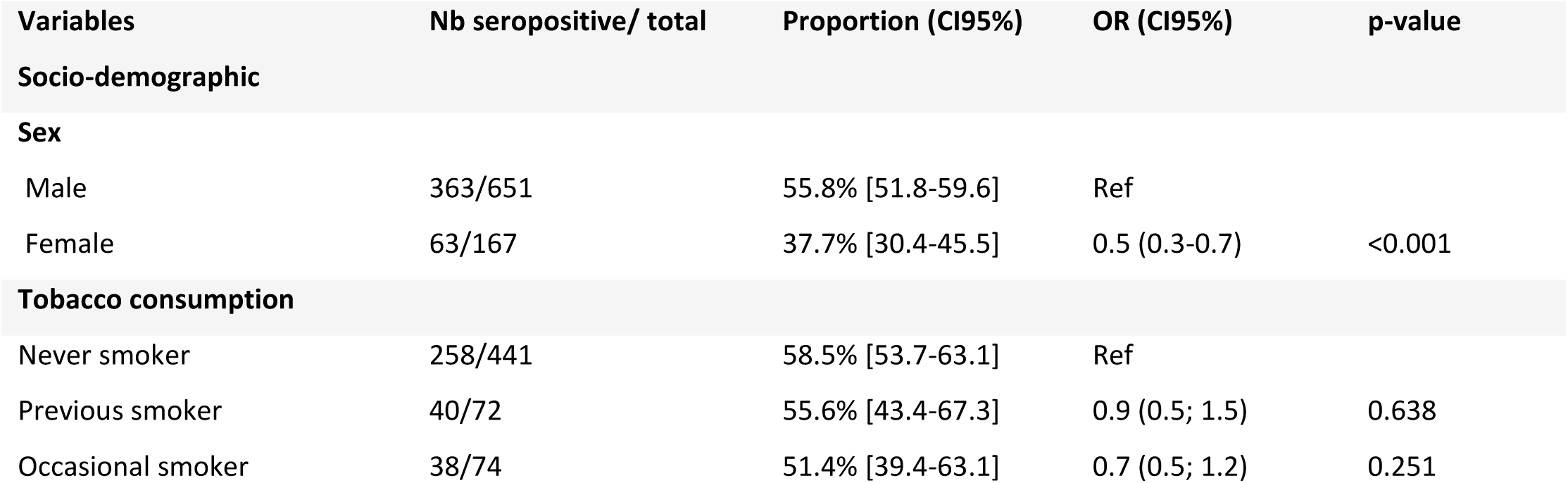
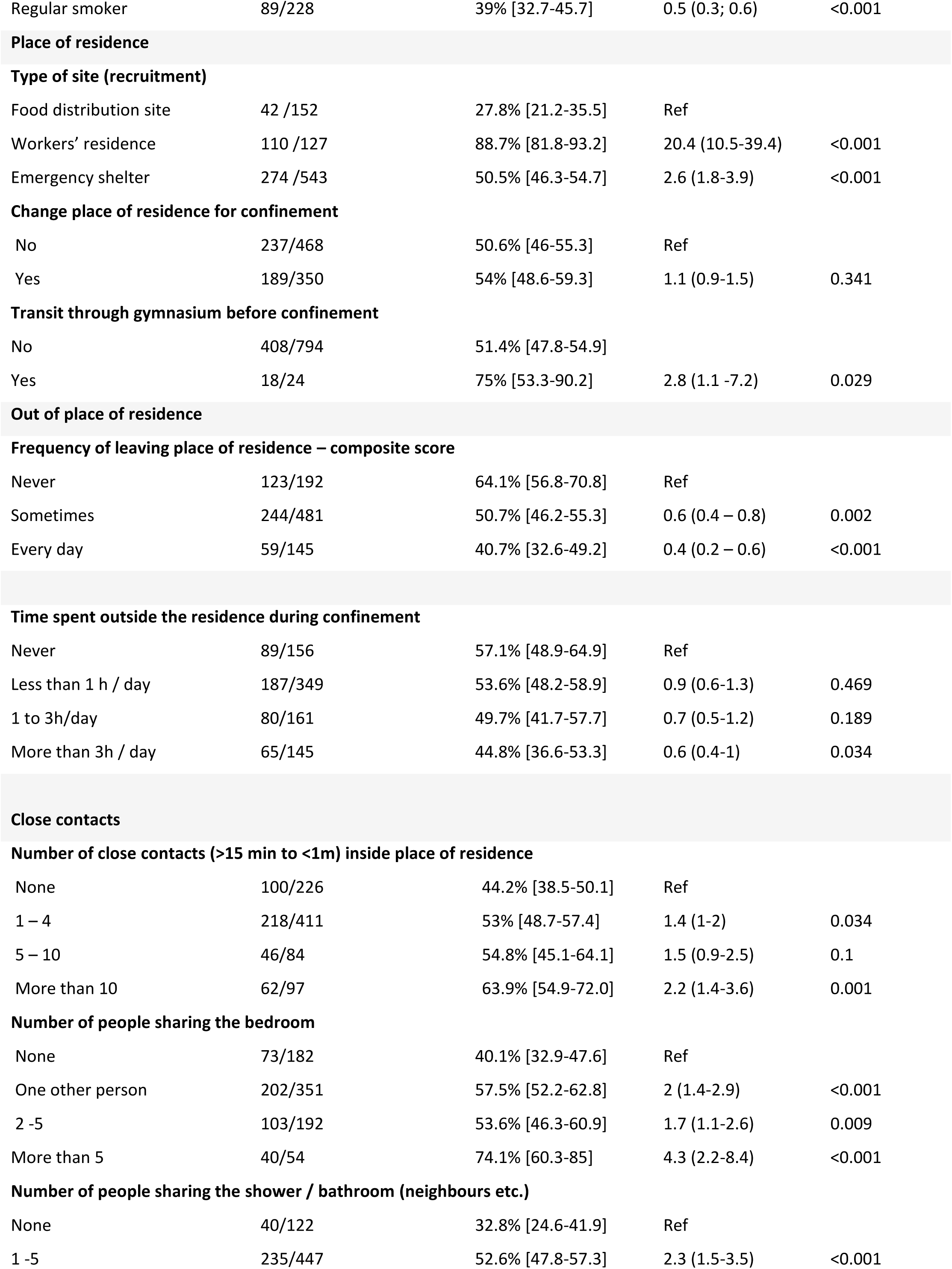
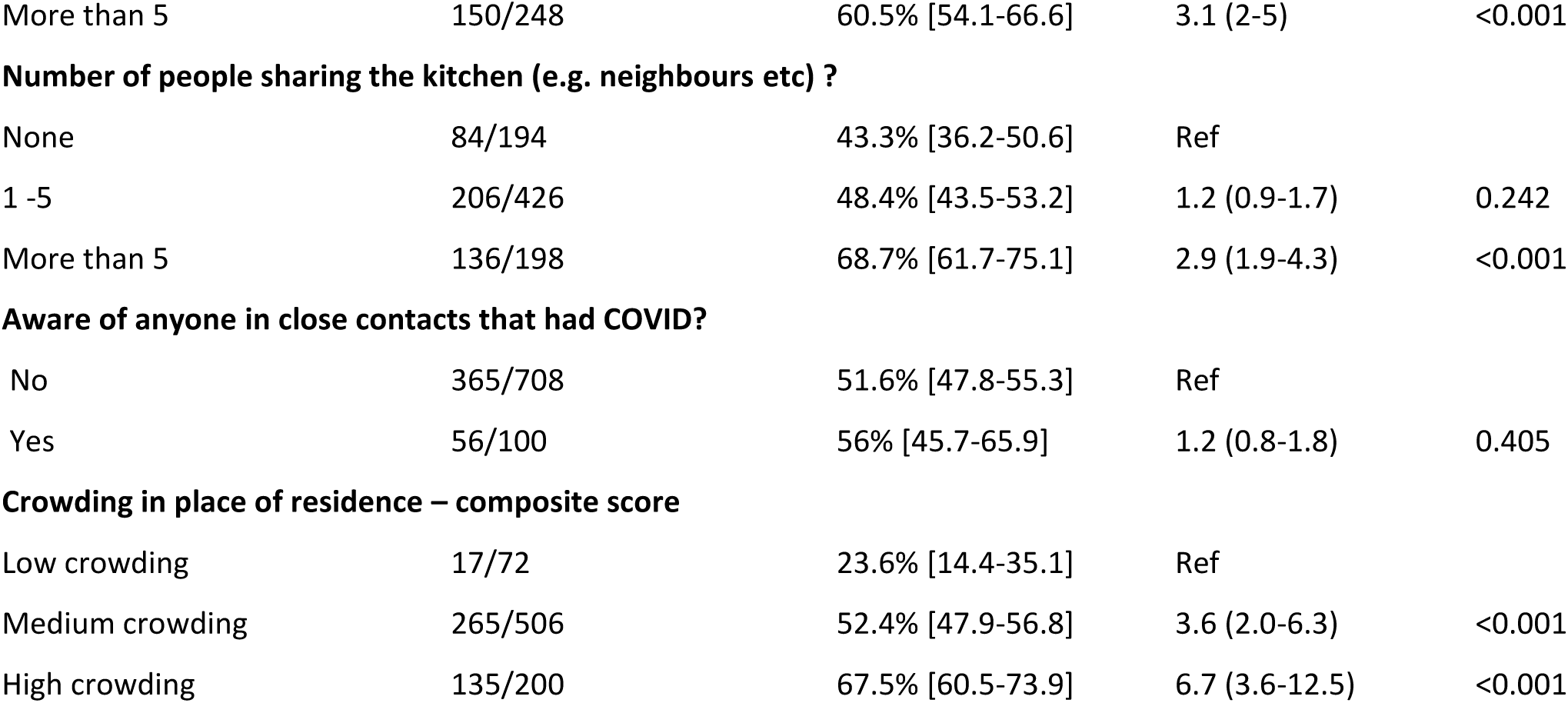
Risk factors for SARS-CoV-2 seropositivity: univariate analysis.

The odds of seropositivity were higher among participants who reported transiting in a gymnasium during the lockdown compared to those who did not (OR 2.8; 95%CI 1.1-7.2; p=0.03). There was no significant difference in seropositivity between individuals who were aware of COVID cases among their close contacts and those who were not aware (OR 1.2; 95%CI 0.8-1.8, p=0.41). The main factor associated with a reduction of exposure to the virus was the frequency of leaving the place of residence: the odds of seropositivity were significantly lower among individuals who left the residence sometimes (OR 0.6, 95%CI 0.4-0.8, p=0.001) or who left the residence nearly every day (OR 0.4, 95%CI 0.2-0.6, p<0.001) compared to those who reported never leaving the residence during the lockdown period. The odds of seropositivity were also lower among individuals who reported regular consumption of tobacco compared to those who reported to have never smoked (OR 0.5, 95%CI 0.3-0.6). However, individuals who reported having adhered to hygiene recommendations most of the time did not have lower odds of seropositivity compared to those who reported not following the recommendations (OR 1.3; 95%CI 0.8-2.1, p=0.27).

In multivariable analysis, the odds of seropositivity remained higher among individuals who lived in more crowded sites (medium: adj. OR 2.7, 95%CI 1.5-5.1, p=0.002; high: adj. OR 3.4, 95%CI 1.7-6.9, p<0.001; compared to individuals with low crowding composite indicators) and those who reported transit through a gymnasium during the lockdown (adj. OR 3.1, 95%CI 1.2-8.1, p<0.001) (Table 2). The association with site type also remained significant, where the odds of seropositivity was 12.0 (adj. OR, 95%CI 5.6-25.6, p<0.001) times higher among individuals from workers’ residences, and 1.7 (adj. OR, 95%CI 1.1-2.7, p=0.025) times higher among individuals from emergency shelters compared to those recruited from food distribution sites. The odds of seropositivity decreased with frequency of leaving the place of residence (left sometimes: adj. OR 0.6, 95%CI 0.4-0.8, p=0.003; left nearly every day: adj. OR 0.4, 95%CI 0.2-0.7, p<0.001; compared to those reporting never leaving the place of residence). Moreover, the odds of seropositivity was lower among regular smokers compared to those who never smoked (adj. OR 0.4, 95%CI 0.3-0.7, p<0.001) and among female than male participants (adj. OR 0.5, 95%CI 0.4-0.8, p=0.01).

**Table 2.** Risk factors for SARS-CoV-2 seropositivity: multivariate analysis.

| Variable | OR | CI 95% | p-value |
| --- | --- | --- | --- |
| **Sex** |  |  |  |
| Male | Ref |  |  |
| Female | 0.5 | 0.4 -0.8 | 0.006 |
| **Frequency of leaving place of residence – composite score** |  |  |  |
| Never | Ref |  |  |
| Sometimes | 0.6 | 0.4 -0.8 | 0.003 |
| Every day | 0.4 | 0.2 -0.7 | 0.001 |
| **Crowding in place of residence – composite score** |  |  |  |
| Low crowding | Ref |  |  |
| Medium crowding | 2.7 | 1.5 -5.1 | 0.001 |
| High crowding | 3.4 | 1.7 -6.9 | <0.001 |
| **Tobacco consumption** |  |  |  |
| Never smoker | Ref |  |  |
| Previous smoker | 0.8 | 0.4 -1.4 | 0.356 |
| Occasional smoker | 0.8 | 0.4 -1.3 | 0.319 |
| Regular smoker | 0.4 | 0.3 -0.7 | <0.001 |
| **Transit through gymnasium before confinement** |  |  |  |
| No | Ref |  |  |
| Yes | 3.1 | 1.2 -8.1 | 0.023 |
| **Type of site (recruitment)** |  |  |  |
| Food distribution site | Ref |  |  |
| Workers' residence | 12.0 | 5.6 -25.6 | <0.001 |
| Emergency shelter | 1.7 | 1.1 -2.7 | 0.025 |

### Symptoms

More than two-thirds of seropositive individuals (68.3%; 95%CI 64.2-72.2) did not report any symptoms during the recall period since 1 March (implying a high proportion of asymptomatic infections). In contrast, 50.2% (95%CI 46.0-54.3) of individuals who did not report symptoms were found to be seropositive. While not statistically significant, there was a trend in association between the report of symptoms associated with COVID and seropositivity: people who were seropositive had a 30% higher odds of reporting symptoms than those who were not (OR 1.3, 95%CI 1.0-1.8, p=0.09). Among the investigated symptoms, six of the twelve reported symptoms were found significantly associated with seropositivity when comparing individuals with severe versus mild or no symptoms (Figure 2), including loss of taste (OR 6.6, 95%CI 2.3-18.9, p<0.001), fever (OR 4.3, 95%CI 2.3-7.8, p<0.001), loss of smell (OR 4.0, 95%CI 1.6-9.9, p=0.003), shivering (OR 3.2, 95%CI 1.5-7.2, p=0.004), fatigue (OR 2.3, 95%CI 1.3-4.2, p=0.007) and cough (OR 2, 95%CI 1.1-3.6, p=0.03). Twenty-five of 238 participants with symptoms (10.5%) reported being tested for COVID previously, of which nine had a positive result and seven were admitted to COVID treatment centres.

**Figure 2.**
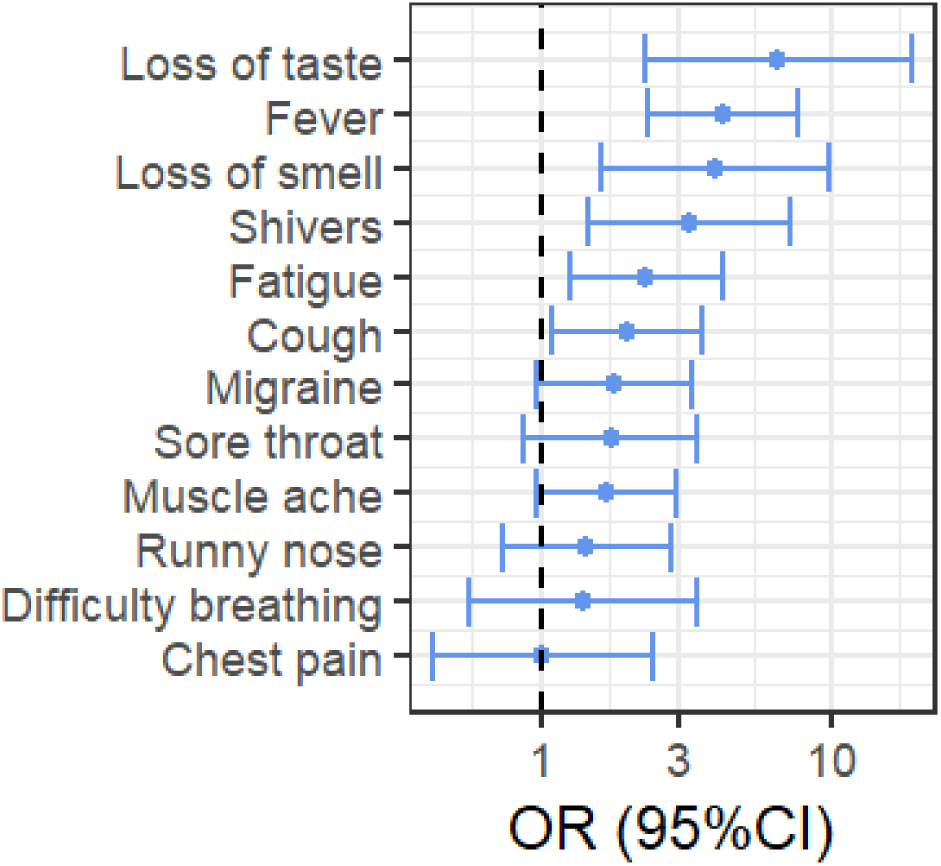
Association of symptoms (strong vs. mild or none) with seropositivity.

## Discussion

This study is the first in Europe to evaluate the exposure to SARS-COV-2 virus of populations who lived in precarious situations during the first months of the COVID-19 pandemic. The results show overall a high exposure to SARS-COV-2 with important variations between sites. This study also identifies overcrowding as the most important factor to explain the variability in exposure rather than reported adherence to the recommended preventive measures. The results further suggest living conditions within the place of residence as the most important risk factor related to exposure, as frequently leaving the place of residence was associated with a lower risk of being seropositive. Coherent with other studies (22,23) evaluating the spectrum of COVID-19 disease, there was a high proportion of asymptomatic or paucisymptomatic infections (up to 68%) in a population that is younger on average than the general European population. Combined with the high proportion of exposed individuals, the high proportion of asymptomatic cases puts into question the pertinence of epidemic surveillance strategies solely based on the identification of symptomatic cases and their contacts.

Our study has several limitations. First, due to the cross-sectional study design, it is not possible to determine when participants became seropositive. In relation to the sampling strategy, the selection of the study sites was not random: the locations were determined by MSF’s operational activities during the first wave of the pandemic in Ile-de-France and other considerations including security constraints and agreement of the sites to give the survey team access. Therefore, our results cannot be extrapolated to other populations living in similarly precarious situations in France or elsewhere. The selection of participants within the study sites could be potentially subject to bias despite the efforts made by the study team to obtain a representative sample; depending on the site, up to one third of those originally selected for inclusion were replaced by another participant. If those replacing the initially selected individuals had a higher risk of exposure, due to, for example, spending more t more time within the place of residence, this could have led to an overestimate of prevalence in the study population. Conversely, refusal to participate could have been higher among those who had previously tested positive, which would bias the seroprevalence estimate in the opposite direction. In addition to possible selection bias, information bias may have affected the measurement of other self-reported exposures,, including living conditions before and during confinement, COVID symptoms and onset period, and adherence to prevention measures. We also cannot exclude social desirability bias, particularly related to compliance with containment and prevention measures, that potentially underestimates exposure. Our efforts to mitigate information bias included the use of standardized pre-tested questionnaires, training of the study team and the participation of translators during interviews.

Previous evaluation of the LuLISA tests have shown very high sensitivity and specificity. These estimates may, however, be revised in the future when results of other evaluation studies become available. Although we cannot exclude misclassification of some tested samples, our sensitivity analysis showed that even assuming a diagnostic test sensitivity or specificity as low as 70%, seroprevalence estimates by type of sites remain high (82.4-95.0% for workers’ residences, 34.7-62.0% for emergency shelters, and 16.2-38.5% for food distribution sites) (Supplementary material).

While several definitions of overcrowding exist, in Europe and North America it is defined as more than one single person or couple per habitable room (24). In this adult study population, only 23.4% [20.4-26.5] overall reported having a single occupancy room; additionally, only 14.9% [12.6-17.6] had their own bathroom and 23.7% [20.8-26.8] their own kitchen. A recent computer-based study of the homeless population in England also emphasized the importance of single-room accommodation, combined with heightened infection prevention methods, in preventing COVID-19 deaths (25). Congregate housing conditions, whether long term or as temporary emergency measures, carry risks during an infectious disease outbreak that should be weighed against the risks in remaining unsheltered. The extent to which these risks can be mitigated, using masks and hand hygiene in crowded living environments and or situations with high seroprevalence as seen in our study, remains uncertain. It may be that the good reported adherence to hygiene measures and frequently reported mask wearing contributed to the high proportion of asymptomatic infections seen in this study (26). Lack of housing is a known risk factor for poor physical and mental health, and overcrowding for infectious diseases (27,28). This poses a complex dilemma for public health policy makers who aim to balance individual health risks and public health in the face of a global pandemic. While ours is the first study to identify overcrowding specifically as a risk factor for SARS-CoV-2 seropositivity among an already vulnerable population in IDF,, large household size has been identified as a risk factor for COVID-19 in a recent study in the general population of France (4) as well as in a study focusing on a cohort of pregnant women tested for SARS-CoV-2 RNA in New York City(29). Many others have shown that the COVID-19 pandemic highlights existing socioeconomic and racial inequalities and inequities (30–32).

In conclusion, the results presented here highlight a high level of exposure to SARS-COV-2 virus, with substantial variability among our sampled population. This underscores the importance of identifying populations at high risk of exposure, understanding the factors that determine this risk to orient prevention and control strategies, and adaptation according to the needs of the affected population. Relocations of individuals living in insecure conditions, particularly those at highest risk of severe disease (i.e. elderly individuals and/ or individuals with certain comorbidities), should be implemented; this process should limit the number of people per room and ensure access to hygienic shared spaces. Based on a high seroprevalence, older average age and other risk factors, adapted strategies for the workers’ residence should be prioritized.

Finally, considering the lack of existing data globally, further epidemiological studies in populations experiencing similar vulnerable conditions should be considered, including qualitative studies, in order to properly protect at-risk populations.

## Data Availability

Databases used for the analyses are available upon request to the corresponding author.

## Acknowledgments

Authors want to thank all the participants to the survey, as well as the managers of the hostels and emergency shelters who made the survey possible.

The authors also thank the team from the Mission France Project Team: Arielle Calmejane, Jean-François Véran, Marianne Viot, Cécile Arondel, Flora Boirin and Vanessa Lalouelle. We also thank the nurses for their hard work and availability: Ophélie Cahu, Aurélie Rawinski, Valérie Tacussel and Elodie Tarral. The interviews have been thoroughly conducted by Rachid Kaddour, Marine Labatut, Saeed Shairzad, Tara Singh, Juliette Dechaux, Margot Dupe, Diane Goulas, Prisla Mogango, Sarah Perretti and Ciamony Mohammadi.

The authors thank Nicolas Escriou, Marion Gransagne (Innovation Laboratory: Vaccine), Stéphane Pètres, François Dejardin (Production and purification of recombinant proteins technological platform), Sébastien Brulé, Sylviane Hoos, Bertrand Raynal and Patrick England (Molecular Biophysics Platform) for the protein target design, production and quality control at Institut Pasteur. The authors thank Olivier Helynck and Hélène Munier-Lehmann for sample handling automation (Chemistry and Biocatalysis, Institut Pasteur). This work was made possible thanks to the financial support obtained through the «URGENCE nouveau coronavirus» fundraising campaign of Institut Pasteur and the financial support of the Fondation Total. LuLISA development has been supported by IARP Pasteur-Carnot MI (2019-2020). We thank the technical support of Berthold France and Berthold Technologies Germany.

## Supplementary Materials

### 1. Further details – serological test results

Figure S1 shows a density plot of observed relative light units per second (RLU/S) among study participants. Positivity for LULISA assays was defined by a cut-off of 10,291 RLU/s; however as the distribution of RLU/s is continuous (and not bimodal); misclassification of participants may occur at values close to the used cut-off. While the main peak in density occurs at RLU/s values lower than the cut-off, q second small peak in density occurs at higher values (50,000-75,000 RLU/s) potentially reflecting individuals with multiple exposures.

**Figure S1.**
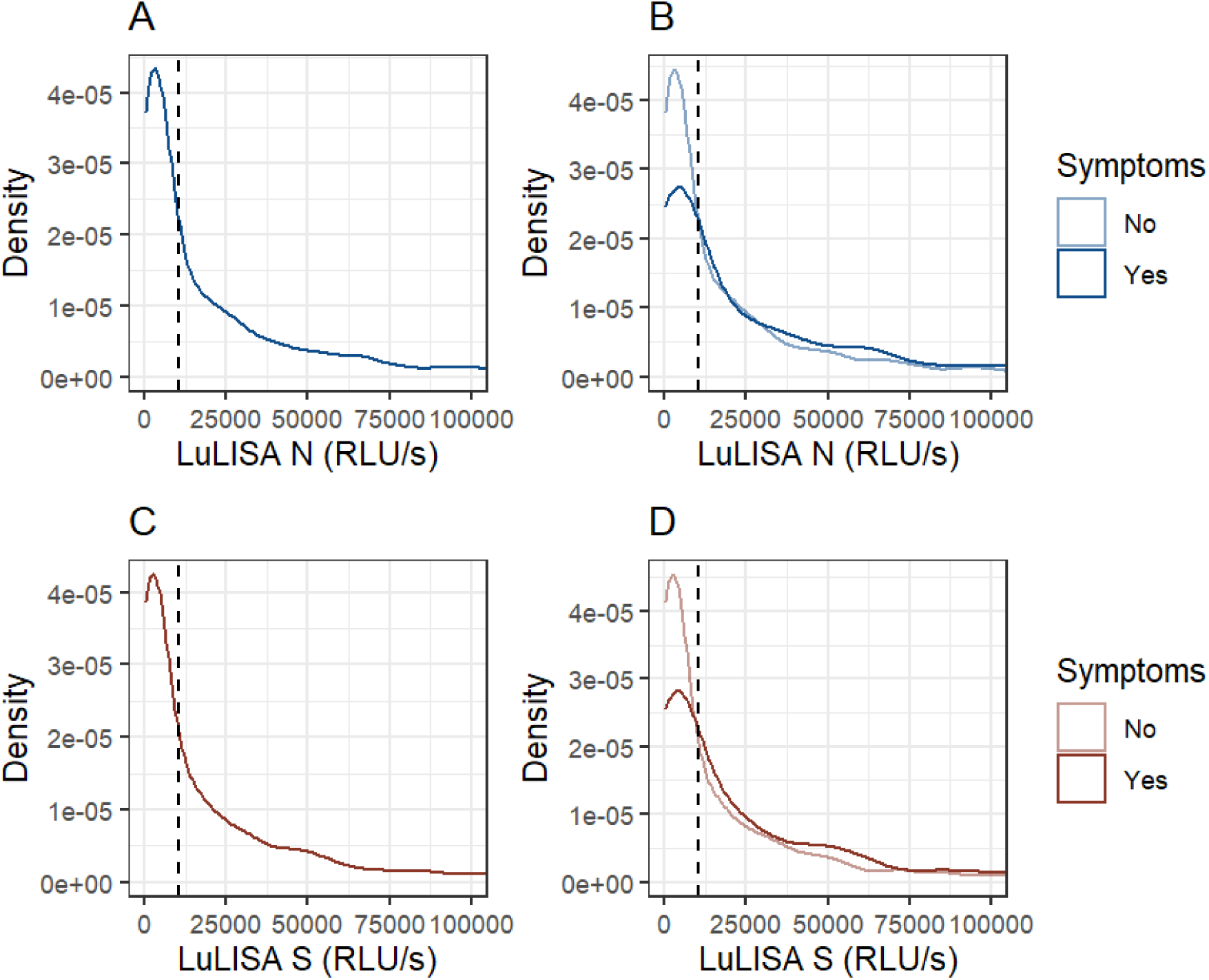
Distribution of LuLISA IgG-N and LuLISA IgG-S RLU/S among study participants. The dashed value indicates the threshold used to define seropositivity (10,291 RLU/s).

The test results by serology technique (LuLISA IgG-N, LuLISA IgG-S, PNT) are summarized in Table S1. PNT can be considered more specific than LuLISA; seroprevalence estimates obtained by PNT can therefore be considered as conservative estimates of seroprevalence. Misclassification based on LuLISA may be related to the transmission setting, where in low prevalence settings seroprevalence estimates by LuLISA may be more strongly overestimated than in high prevalence settings, which is reflected by a higher relative prevalence of LuLISA compared to PNT. This tendency is also visible in the relative prevalences of LuLISA compared to PNT by recruitment sites (Figure S2). Overall, concordance between the techniques was pretty strong (Figure S3).

**Table S1.**
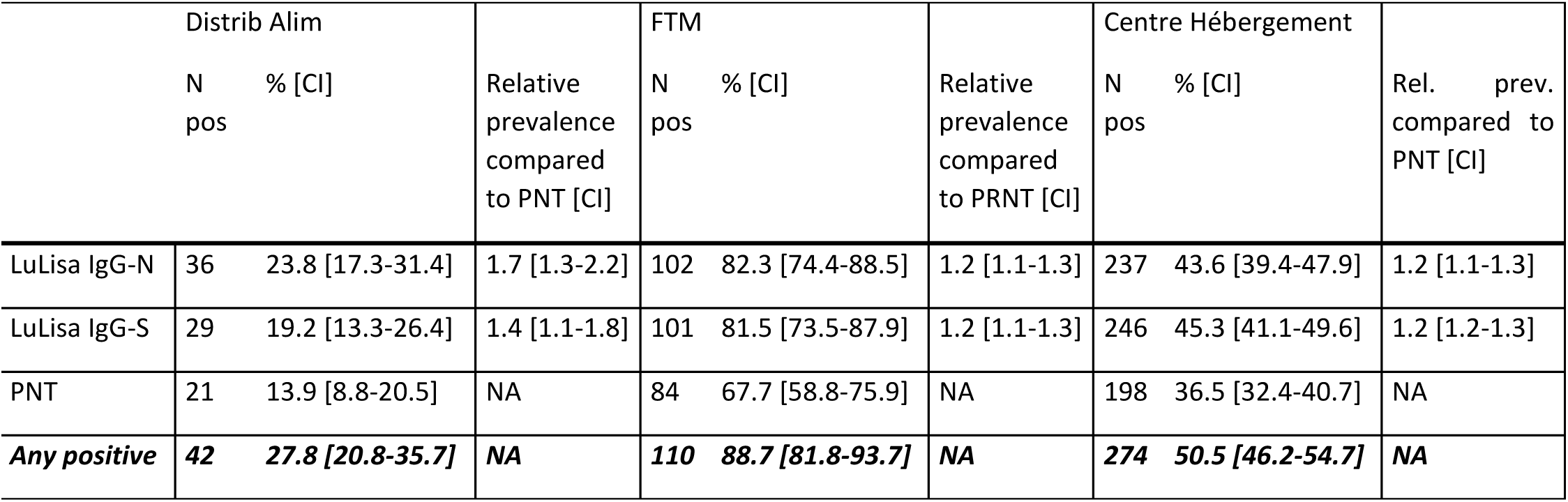
Seropositivity by type of test.

**Figure S2.**
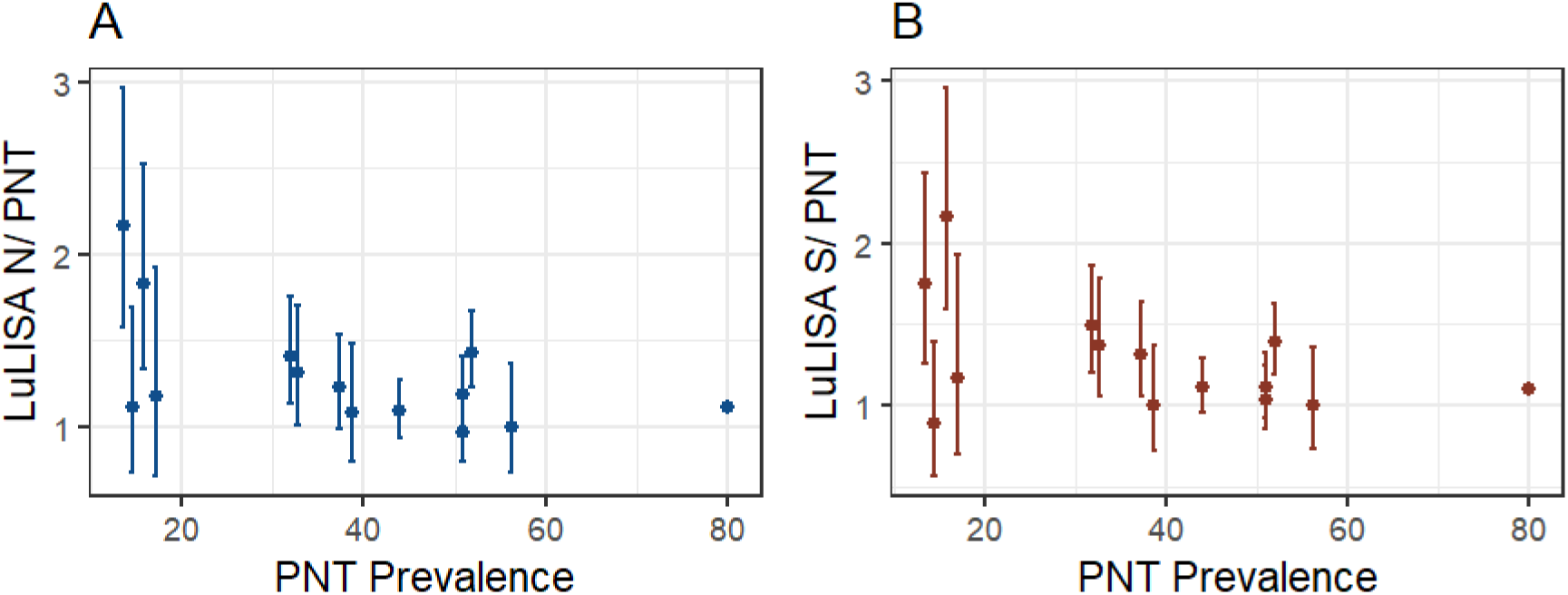
Relative prevalence of LuLISA IgG-N and LuLISA IgG-S compared to the prevalence estimated by PNT. For the site with the highest prevalence, confidence intervals could not be estimated due to failure of model convergence.

**Figure S3.**
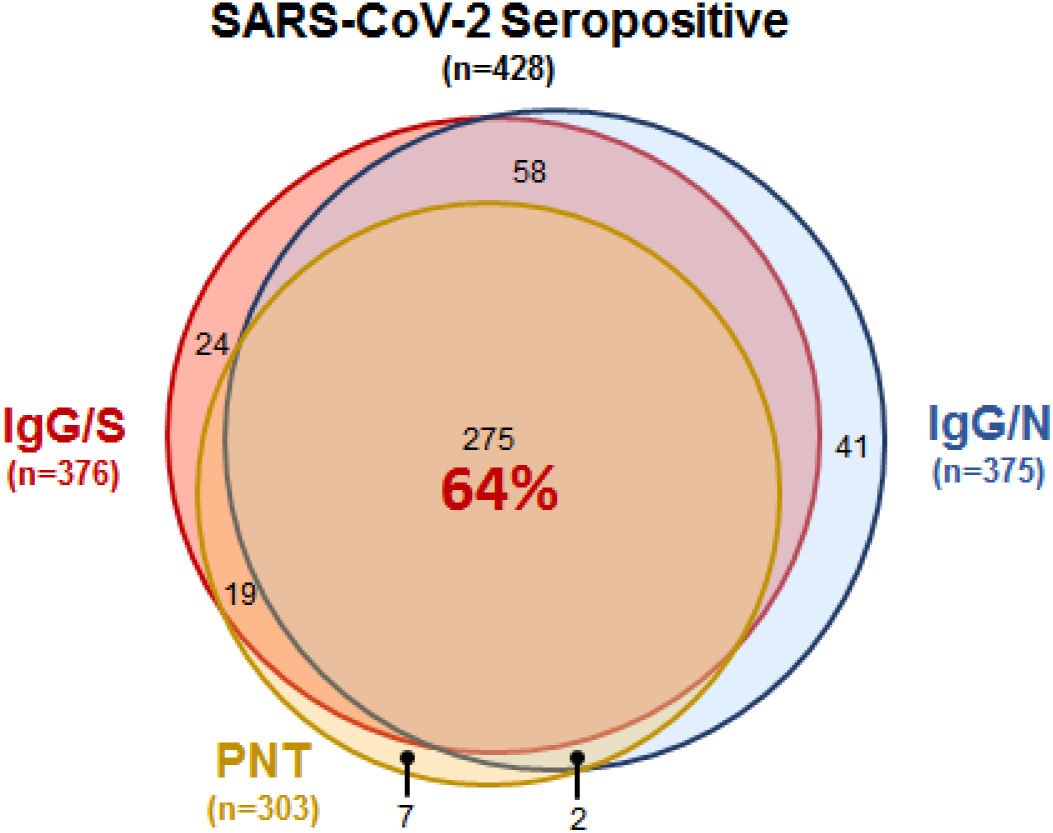
Seropositivity according to the 3 different techniques. Overlap shows that 64% of the 428 seropositives (as defined per protocol) are captured by all 3 serology techniques, showing a strong concordance.

### 2. Composite crowding indicator

We first categorized number of people sharing the room (0, 1, 2-5, and >5 people), sharing a sanitary facility (0, 1-5, and >5 people), sharing a kitchen (0, 1-5, and >5 people), and number of close contacts per day (0, 1-4, 5-10, >10 people). We then created a composite indicator of the sum of levels of the categorical variable and created categories as low (values ≤5), medium (values 6-9), and high (values ≥10). For example, an individual sharing the room with no other person (level 1), the kitchen and sanitary facilities with 1 other person (each of level 2), and who reported on average 1 close contact per day (level 2) would be assigned an indicator value of seven – representing the medium crowding category.

### 3. Sensitivity analysis – seroprevalence estimates by type of sites

We performed a sensitivity analysis of seroprevalence estimates taking uncertainty about diagnostic test performances into account as described in “Seroprevalence of anti-SARS-CoV-2 IgG antibodies in Geneva, Switzerland (SEROCoV-POP): a population-based study. https://doi.org/10.1016/S0140-6736(20)31304-0”. To estimate seroprevalence and 95% credible intervals (95% CrI) of site types we used a Bayesian logistic regression model including the type of site as fixed effect, integrated with a binomial model of assumed diagnostic test sensitivity and specificity.

We then made several assumptions about diagnostic test sensitivities as evaluated:

35 true positive samples out of 35 positive controls (Sens 35/35) and 32 true negative samples out of 32 negative controls (Spec 32/32) (as reported in “Sensitivity in Detection of Antibodies to Nucleocapsid and Spike Proteins of Severe Acute Respiratory Syndrome Coronavirus 2 in Patients With Coronavirus Disease 2019. https://doi.org/10.1093/infdis/jiaa273”)

i. 70 true positive samples out of 100 positive controls (Sens 70/100) and 70 true negative samples out of 100 negative controls (Spec 70/100)
ii. 100 true positive samples out of 100 positive controls (Sens 100/100) and 70 true negative samples out of 100 negative controls (Spec 70/100)
iii. 70 true positive samples out of 100 positive controls (Sens 70/100) and 100 true negative samples out of 100 negative controls (Spec 100/100)

The estimated seroprevalence for emergency shelters ranged from 34.7-62.0% (compared to the main estimate of 50.5%); for food distribution sites it ranged from 16.2-38.5% (compared to the main estimate of 27.8%); and for workers’ residences it ranged from 82.4-95.0% (compared to the main estimate of 88.7%).

**Table S2.** Seroprevalence estimates based on different assumptions about diagnostic test performances. Main estimates are based on the prevalence estimated from the data and 95%CI based on the Clopper-Pearson method.

| Site type | Main Estimates | Sens 35/35; Spec 32/32 | Sens 70/100; Spec 70/100 | Sens 100/100; Spec 70/100 | Sens 70/100; Spec 100/100 |
| --- | --- | --- | --- | --- | --- |
|  | % (95%CI) | Median (95%CrI) | Median (95%CrI) | Median (95%CrI) | Median (95%CrI) |
| Emergency shelters | 50.5% [46.3-54.7] | 50.1% [44.5-55.5] | 45.8% [34.4-56.4] | 34.7% [25.7-42.3] | 62.0% [55.6-69.3] |
| Food distribution sites | 27.8% [21.2-35.5] | 29.7% [22.1-37.4] | 22.8% [12.9-34.7] | 16.2% [9.0-25.1] | 38.5% [29.7-48.2] |
| Workers' hostels | 88.7% [81.8-93.2] | 87.7% [80.6-93.6] | 92.1% [84.5-97.1] | 82.4% [72.9-89.6] | 95.0% [89.3-98.2] |

### 4. Sensitivity analysis on model selection – multivariable analysis of risk factors

Here we present results of a sensitivity analysis of the multivariable analysis of risk factors for seropositivity. In addition to the main model which had the lowest Akaike Information Criterion (AIC) of 938-we additionally tested models without site random effects, without adjusting for the type of site, or with number of people sharing the room as alternative crowding indicator.

**Model 2: Including a random effect for sites but not adjusting for the type of site – crowding indicator: cumulative crowding indicator**

**Table S3.**
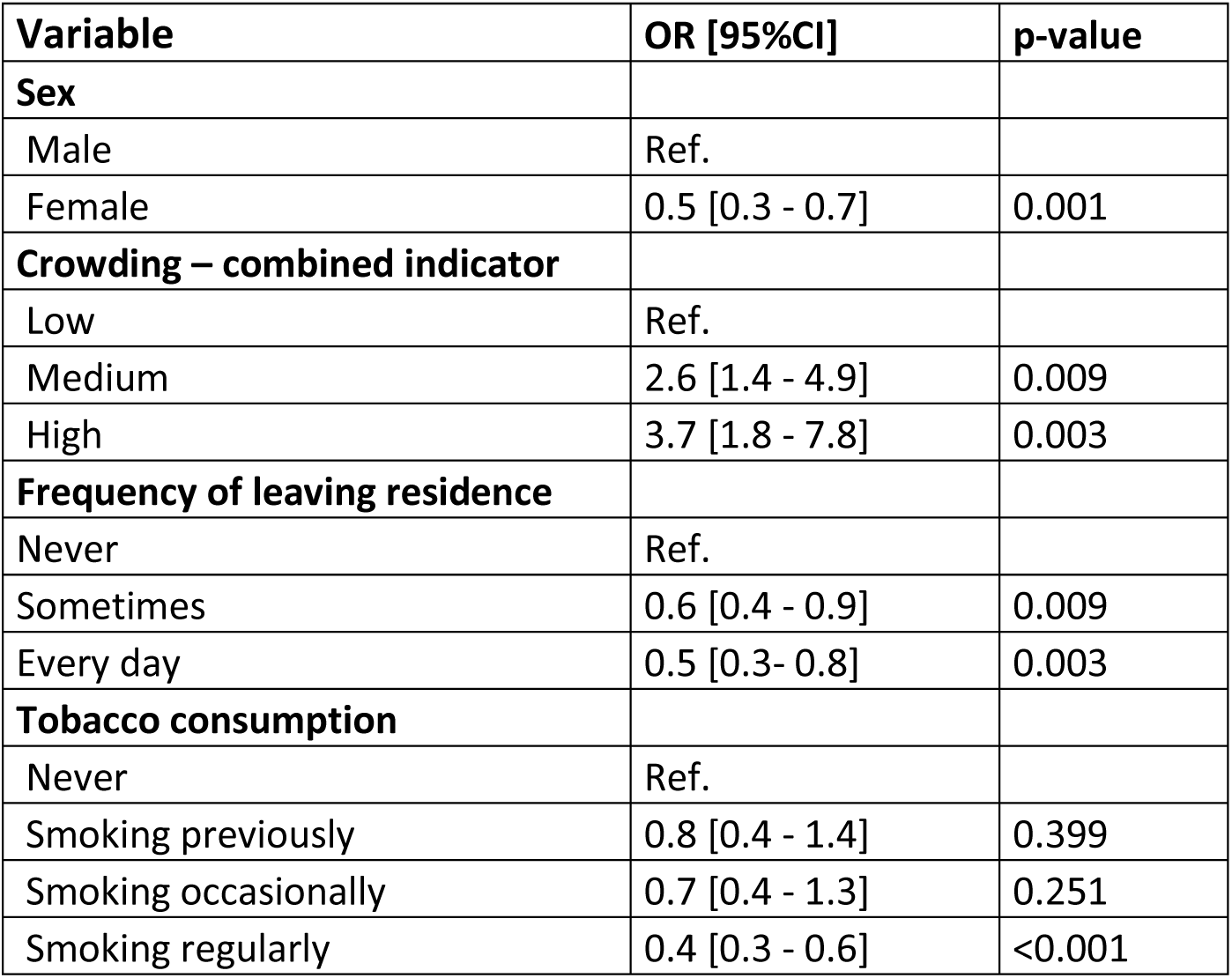
Model 2. Akaike Information Criterion (AIC) 955.

**Model 3: Including a random effect for sites but not adjusting for the type of site – crowding indicator: number of people sharing room**

**Table S4.**
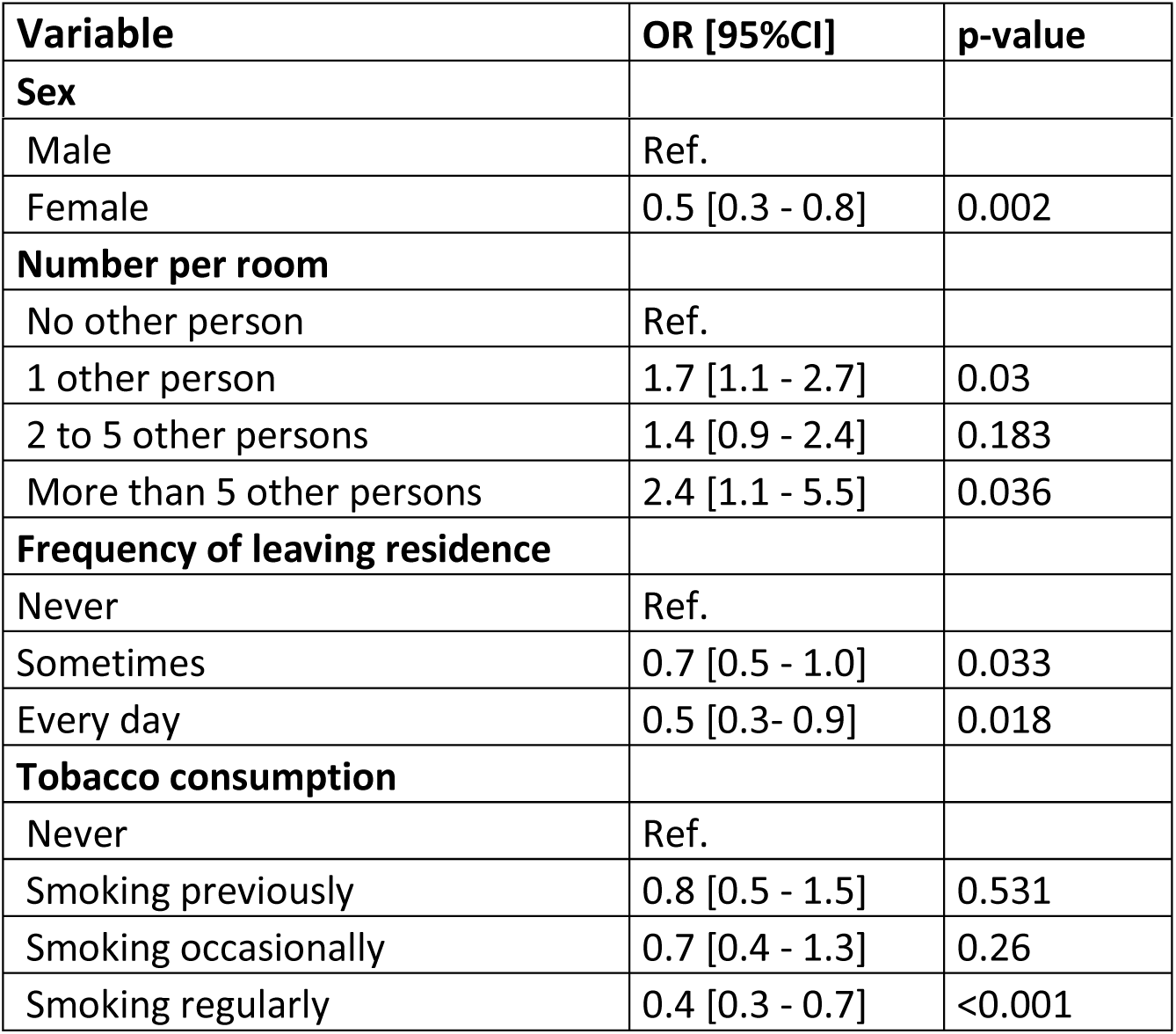
Model 3. Akaike Information Criterion (AIC) 965.

**Model 4: No random effect for sites and not adjusting for the type of site – crowding indicator: cumulative crowding indicator**

**Table S5.**
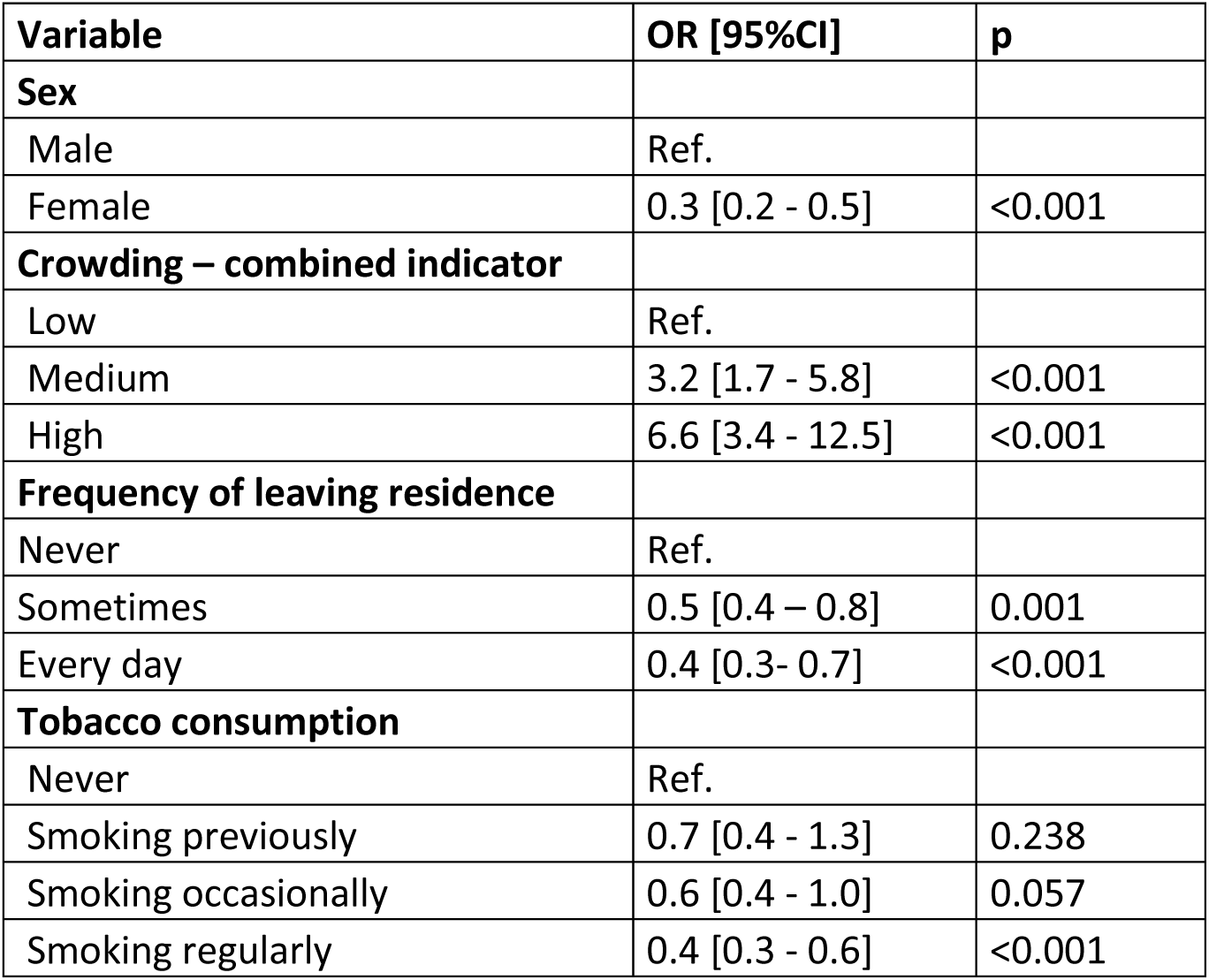
Model 4. Akaike Information Criterion (AIC) 984.

| Variable | OR [95%CI] | p |
| --- | --- | --- |
| **Sex** |  |  |
| Male | Ref. |  |
| Female | 0.3 [0.2 - 0.5] | <0.001 |
| **Crowding – combined indicator** |  |  |
| Low | Ref. |  |
| Medium | 3.2 [1.7 - 5.8] | <0.001 |
| High | 6.6 [3.4 - 12.5] | <0.001 |
| **Frequency of leaving residence** |  |  |
| Never | Ref. |  |
| Sometimes | 0.5 [0.4 – 0.8] | 0.001 |
| Every day | 0.4 [0.3- 0.7] | <0.001 |
| **Tobacco consumption** |  |  |
| Never | Ref. |  |
| Smoking previously | 0.7 [0.4 - 1.3] | 0.238 |
| Smoking occasionally | 0.6 [0.4 - 1.0] | 0.057 |
| Smoking regularly | 0.4 [0.3 - 0.6] | <0.001 |

## Notes

### Competing Interest Statement

The authors have declared no competing interest.

### Clinical Trial

Study was registered in France only, it is not a clinical trial, but an epidemiological survey in population.

### Funding Statement

No external funding was received

### Author Declarations

This protocol was approved by the MSF ethics review board on 18 June 2020, reference number 2044) and by the Comite de Protection des Personnes (CPP), Ile de France, Paris XI, approved 19 June 2020 (reference number 20050-62628).

